# A Study on Antibiotic Susceptibility Pattern of DNase Positive Staphylococci Isolated from Selected Libraries of Halls of Residence in Obafemi Awolowo University, Ile-Ife, Osun State, Nigeria

**DOI:** 10.1101/2025.06.29.25330498

**Authors:** Victor Olamiposi Olaiya

## Abstract

**Introduction:** DNase-positive staphylococci are clinically significant pathogens capable of causing community-acquired infections. This study assessed their prevalence and antibiotic resistance patterns on reading tables in student halls of residence at Obafemi Awolowo University, Ile-Ife, Nigeria.

**Materials and Methods:** Sixty-eight tables across four halls were swabbed. Isolates were identified as presumptive staphylococci via Gram staining (cocci in clusters), catalase positivity, and oxidase negativity. DNase testing confirmed 12 isolates from 23 staphylococci. Kirby-Bauer disc diffusion tested susceptibility to seven antibiotics (rifampicin, cefoxitin, erythromycin, clindamycin, chloramphenicol, tetracycline, ciprofloxacin) using CLSI (2016) standards.

**Results:** Among DNase-positive isolates, resistance rates were: tetracycline (42%, 5/12), chloramphenicol (33%, 4/12), erythromycin (17%, 2/12), and rifampicin/cefoxitin/clindamycin (8% each, 1/12). All isolates (100%) were susceptible to ciprofloxacin.

**Discussion/Conclusion:** Reading tables serve as reservoirs for antibiotic-resistant staphylococci, linked to inadequate cleaning. Ciprofloxacin showed maximal efficacy, but judicious use is advised. Regular surface disinfection is critical to prevent community transmission in academic environments.

## 1. Introduction

### 1.1 Fomites

Fomites are known as inanimate objects, and can carry infectious microorganisms. Fomites are potential reservoir in the transmission of pathogen either directly, by surface-to-mouth contact, or indirectly, by contamination of fingers and subsequent hand-to-mouth, hand-to-eye or hand-to-nose contact (Haas *et al.,* 1999; Nicas and Sun, 2006). Fomites include tables, door knobs, identification cards, writing materials and clothing materials. Body fluids such as mucus, saliva, nasal secretions, blood, urine, and faeces may contain potential pathogens that can be transmitted by fomites to a susceptible individual (Baker *et al.,* 2001; Baker *et al.,* 2004). Enteric and respiratory pathogens can survive on fomites for extended periods, ranging from hours to months. Their range of survival depends on the type of organism, the number of organisms deposited, and the variable environmental conditions. Contaminated hands play a critical role as a route of exposure (Abad *et al.,* 2001; Kramer *et al.,* 2006; Boone and Gerba 2007). Fomites come in contact with the body, and examples of these fomites are wristbands, necktie, tables, lanyards, and head bands. Microorganisms can survive on these surfaces for a very long time (Kramer *et al.,* 2006). This situation might lead to opportunistic pathogenicity, which could cause infection in susceptible hosts (Grice and Segre, 2011; Wilson, 2005).

### 1.2 Microorganisms and Antibiotic Resistance

Microorganisms are universal, and diverse microbes are often transferred to everyday objects from the environment and infected individuals. Microorganisms are transmissible by means of air, fomites (which incorporate tables, knives, mobile phones, identity cards or seats), skin, food, water and other relational contacts, and in several cases, they can cause contaminations which lead to diseases (Mercola, 2000; CDC, 2003). Resistance of pathogenic organisms to antibiotics has become a worldwide problem with serious consequences on the treatment of infectious diseases. The increase in use/abuse of antibiotics in human medicine, agriculture and veterinary is primarily contributing to this phenomenon. Examples of multidrug resistant pathogens include, *Escherichia coli*, *Klebsiella pneumoniae*, *Acinetobacter baumannii,* penicillin-resistant *Streptococcus pneumoniae*, vancomycin-resistant *Enterococcus,* extensively drug-resistant *Mycobacterium tuberculosis* and methicillin-resistant *Staphylococcus aureus* (Goffin and Ghuysen, 1998; Alekshun and Levy, 2007).

### 1.3 Staphylococci

Staphylococci are Gram-positive cocci, and are in the order bacillales. The term “*Staphylococcus*” was coined from the Greek word “staphyle,” meaning bunch of grapes, for their ability to form small grape-like bunches, and the term “coccus,” meaning grain or berry. The genus *Staphylococcus* is composed of Gram-positive bacteria with diameters of 0.5-1.5 µm, characterized by individual cocci that divide in more than one plane. These microscopic organisms are non-motile, non-spore formers, facultative anaerobes, featuring a complex nutritional requirement for growth, a low G+C content of DNA (in the range of 30-40 mol%), a resilience to high concentrations of salt and resistance to heat (Madigan and Martinko, 2005; Giorgio *et al.,* 2015).

Amidst the genus *Staphylococcus,* they can cause a wide scope of infections which includes, impetigo, folliculitis, superficial and deep skin abscesses. It also causes wound infections, osteomyelitis, suppurative arthritis, pneumonia, pleural emphysema, meningitis, septicaemia, and endocarditis (Von Eiff *et al.,* 2001; Von Eiff *et al.,* 2002; Tarazona *et al.,* 2007; Sakine *et al.,* 2007).

#### 1.3.1 Microbiological Differentiation Of Staphylococci

##### 1.3.1.1 Growth Under Various Conditions

Staphylococci are facultative anaerobes as they grow most rapidly under aerobic conditions, and in the presence of carbon dioxide (CO_2_). The colonies of *S. aureus* are β-haemolytic due to the production of few haemolysins: α-toxin, β-toxin, γ-toxin, and δ-toxin. Some *S. epidermidis* strains are β-haemolytic due to the production of δ-toxin. (Tegmark *et al.,* 1998). Pigmentation is more pronounced after 24 hours and when held at room temperature, or in media enriched with acetate or glycerol monophosphate (Martin *et al.,* 2000; Amos *et al.,* 2006).

The pigments are not produced in anaerobic conditions or by small colony variants (Proctor *et al.,* 2002). The formation of pigments depends on the, stress sigma factor, σ^B^ (Katzif *et al.,* 2005). Staphylococci can grow in a wide pH range of 4.8–9.4, able to resist drying, and can survive at high temperature of 60°C for 30 minutes. In addition, *S. aureus* can grow in high-salt medium due to its production of osmo-protectants (Nester *et al.,* 2004), and it can tolerate 7.5–10% sodium chloride (NaCl). The ability of *S. aureus* to ferment mannitol is the basis for differentiating it from *S. epidermidis* and *S. saprophyticus*. When grown on mannitol salt agar, fermentation of mannitol creates a yellow zone around the colony. In addition to mannitol, *S. aureus* can use and metabolize glucose, xylose, lactose, sucrose, maltose, and glycerol. Further differentiation of staphylococci can be achieved by growth in the presence of novobiocin (Vickers *et al.,* 2007).

##### 1.3.1.2 Other Methods For Differentiation Of Stapylococci

A 12^th^ group of *Staphylococcus species* (*Staphylococcus caseolyticus*) have been moved to a new genus, *Macrococcus,* which differs from *Staphylococcus* based on the susceptibility of *Staphylococcus* to lysis by lysostaphin and the oxidase test. (Geary and Stephens, 1986; Kloss *et al.,* 1998). Lysostaphin is a metalloendopeptidase that targets the pentaglycine bridge of peptidoglycan (Grundling and Scheewind, 2006). Phenol soluble modulins have been associated with more severe staphylococcal infections, and require specialized chromatography and mass spectrometry for identification and quantification (Klingenberg *et al.,* 2007). Polymerase chain reaction (PCR) testing has been used, and its use is increasing rapidly research laboratories. One of the most reliable PCR tests for *S. aureus* detects the presence of the thermo nuclease gene *nuc* (Becker *et al.,* 1998; Becker *et al.,* 2001; Becker *et al.,* 2005). PCR can likewise be used to test for the presence of genes encoding Panton-Valentine leucocidin (PVL), which is indicative of strains associated with community acquired methicillin-resistant *S. aureus* (CA-MRSA) (Finck-Barbancom *et al.,* 1993).

### 1.4 Classification Of Staphylococci

Domain: Bacteria

Kingdom: Bacteria

Phylum: Firmicutes

Class: Bacilli

Order: Bacillales

Family: Staphylococcaceae

Genus: *Staphylococcus*

The taxonomy is based on the 16s RNA sequences and most of the species fall into 11 clusters (Takahashi *et al.,* 1999).

***Staphylococcus aureus*** **group** – *S. aureus, S. simiae*

***Staphylococcus auricularis*** **group** – *S. auricularis*

***Staphylococcus carnosus* group –** *S. carnosus, S. condiment, S. massilliensis, S. piscifermentans, S. simulans*

***Staphylococcus epidermidis* group –** *S. captitis, S. caprae, S. epudermidis, S. saccharolyticus*

***Staphylococcus haemolyticus* group –** *S. haemolyticus S. devriesei, S. hominis*

***Staphylococcus lugduensis* group** – *S. lugdunensis*

***Staphylococcus saprophyticus* group –** *S. arlettae, S. cohnii, S. equorum, S. gallinarum, S. kloosii, S. leei, S. nepalensis, S. saprophyticus, S.succinus, S.xylosus*

***Staphylococcus sciuri* group** -*S. fleurettii, S. lentus, S. sciuri, S. stepanovicii, S. vitulinus*

***Staphylococcus simulans* group** – *S. simulans*

***Staphylococcus warneri* group** – *S. pasteuri, S. warneri*

***Staphylococcus hyicus-intermedius* group** – *S. agnetis, S. chromogenes, S.felis, S. delphini, S. hyicus, S. intermidis S. lutrae, S. microtti, S. muscae, S. pseudintermedius, S. rostri, S. schleiferi.* (Takahashi *et al.,* 1999).

Staphylococcal species have been separated using distinctive biochemical tests, for example, coagulase test and the DNase test (Martineau *et al.,* 2000). Organisms presumptively identified as *Staphylococcus aureus* can be identified based on DNase tests and growth on mannitol salt agar (MSA*). Staphylococcus aureus* gives a positive coagulase and DNase test while most species of coagulase negative staphylococci also test negative to DNase test reaction (Kateete *et al.,* 2010).

### 1.5 Dnase Positive Staphylococci

DNase positive staphylococci are immune-suppressing pathogens which could be part of the diverse groups microorganisms that may present on fomites in the environment, they have been classified based on their catabolic action on the DNA fragmenting in the DNase agar plate which is reflective in the formation of clear zone around the line streak after the addition of one normal (1N) hydrochloric acid. DNase positive staphylococci usually show positive result to the coagulase test. This test is use in differentiating *Staphylococcus aureus* from *Staphylococcus epidermidis,* as the former is DNase positive while the latter is DNase negative. Species that are slightly positive for DNase reaction include: *Staphylococcus capitis* (Pratiksha, 2015). The zymogen granules that are present within the bacteria is a reservoir of many families of the deoxyribonuclease enzyme, which can be found in the nuclear envelope of the pathogen (Buchana *et al.,* 2006). The enzyme deoxyribonuclease is entirely responsible for endonucleolytic hydrolysis of deoxyribonucleic acid (Meena and Roy, 2006).

#### 1.5.1 Staphylococcus Aureus Infection

*Staphylococcus* was first discovered in 1880 in Aberdeen, Scotland, by the surgeon Sir Alexander Ogston in the discharge from a surgical ulcer in a knee joint (Ogston, 1984). This was later named *Staphylococcus aureus* by Friedrich Julius Rosenbach (www.whonamedit.com.). *S. aureus* are part of the normal flora of the body and lower reproductive tract of women (Kluytmans *et al.,* 1997; Cole *et al.,* 2001; Senok *et al.,* 2009). *S. aureus* can cause a wide range of illness from the minor skin infections, for example, pimples, impetigo bubbles, cellulitis, folliculitis, burnt skin disorder and carbuncles (Girolomoni *et al.,* 2016) to the major infections, for example, pneumonia, meningitis, osteomyelitis, endocarditis, bacteraemia, and sepsis (Bethesda, 2004).

##### 1.5.1.1 *Staphylococcus Aureus* Infection Treatment

Staphylococcal infections may range from infections of neonates to adults. The best treatment for staphylococcal infection in humans is antimicrobial exposure at the site of infection or surgical control of the infection. For antimicrobial therapy of *S. aureus* infections, it should be classified into methicillin susceptible *S. aureus* (MSSA) and methicillin resistant *S. aureus* (MRSA), further classification should be made between the more antibiotic-resistant hospital acquired strains (HA-MRSA) and community-acquired strains (CA-MRSA). MRSA strains cannot be eliminated by ampicillin or penicillin and β-lactamase-stable antistaphylococcal penicillins (oxacillin, dicloxacillin amongst others). CA-MRSA do not carry a resistance profile that is usually seen in HA-MRSA strains which carry relatively large antibiotics resistance genes cassettes with concurrent resistance to clindamycin, macrolides (erythromycin, azithromycin, etc.), and aminoglycosides (Bubeck *et al.,* 2008).

### 1.6 Antibiotics

Antibiotics are molecules that kill, or stop the growth and development of microorganisms, including both bacteria and fungi. Antibiotics that kill bacteria are called "bactericidal". Antibiotics that stop the growth of bacteria are called "bacteriostatic” (Thenmozhi *et al.,* 2014). Thus, antibiotics have been classified traditionally as either bactericidal or bacteriostatic (Carter *et al.,* 1995).

### 1.7 Classes Of Antibiotics

Antibiotics are classified based on:

a. The producing micro-organisms (Source of production)
b. Activity spectrum (wide activity spectrum or narrow activity spectrum)
c. Metabolic pathways of biosynthesis
d. Chemical structure

Antibiotics have been classified based on general similarity of chemical structure.

1. Penicillin and related antibiotics: All members of this group have a β-lactam ring in their structure. This group includes the natural penicillins, the semisynthetic penicillins and cephalosporins. Members of penicillin class include penicillin G, penicillin V, oxacillin (dicloxacillin), methicillin, nafcillin, ampicillin, amoxicillin, carbenicillin, piperacillin, mezlocillin, and ticarcillin (Boundless, 2016).
2. Aminoglycoside antibiotics: The streptomycin was the first drug discovered in this class of antibiotics in 1943 (Mahajan and Balachandran, 2012). All members of this group have amino sugars in glycoside linkage. This group comprises the streptomycin, neomycin, kanamycin, paromomycin gentamycin, tobramycin and amikacin (Peterson, 2008; Talaro and Chess, 2008).
3. Macrolide antibiotics: All these consist of a macro cyclic lactone ring to which sugars are attached. This group includes erythromycin, oleandomycin and puromycin (Moore, 2015).
4. Tetracycline antibiotics: The Tetracyclines are derivatives of the poly cyclic naphthalene carboxamide. This group consists of tetracycline, chlortetracycline, demeclocycline, oxytetracycline and minocycline (Fuoco, 2012).
5. Chloramphenicol: This antibiotic is in a class in itself. It is a nitrobenzene derivative of dichloroacetic acid.
6. Peptide antibiotics: These antibiotics form a large group but very few have found therapeutic application. These antibiotics are composed of peptide-linked amino acids which includes both D and L-forms. Antibiotics in this category include bacitracin, gramicidin and the polymyxins. (Allen and Nikas, 2003)
7. Antifungal antibiotics: This group has two main sub-groups

a. polyenes which have a large ring with a conjugated double bond system. In this group, most important antibiotics are hystain and amphotericin B,
b. the other group including 5-fluro cytosine, clotrimazole and Griseofulvin (Stawinski *et al*., 2013; Xu *et al.,* 2014).

Unclassified: These antibiotics have varying structures. They are not classified among the main groups described above, antibiotics in this group include cycloserine, fusidic acid, novobiocin, prasinnomycin, spectinomycin and vancomycin (Power and Diginawala, 2001).

In preparation, antibiotics are subdivided into the following seven groups.

1. Penicillins (including semi synthetic methicillin, oxacillin, ampicillin) and cephalosporins.
2. Broad-Spectrum antibiotics {tetracycline and their derivatives).
3. Streptomycin group (streptomycin, neomycin).
4. Reverse antibiotics (erythromycin, chloramphenicol ristomycin, novobiocin).
5. Antifungal (Levorin, nystatin).
6. Antituberculous (streptomycin, kanamycin, pirlimycin).
7. Antineoplastic which includes: bruneomycin, olivomycin (Power and Diginawala, 2001).

### 1.8 How Do Antibiotics Work?

Antibiotics have been known to have five (5) major modes of action and activity against microorganisms, and they include,

#### 1.8.1 Interference With Cell Wall Synthesis

Beta-lactam antibiotics like penicillin and cephalosporin hinder proteins and enzymes that oversee the development of peptidoglycan layer (Benton *et al.,* 2007).

#### 1.8.2 Inhibition Of Protein Synthesis

Oxazolidinones, one of the new class of antibiotics, interface with the A-site of the bacterial ribosome where they should interfere with the placement of the aminoacyl-tRNA. Tetracyclines interfere with protein synthesis by binding to 30S subunit of ribosome, thereby weakening the ribosome-tRNA interaction. Macrolides bind to the 50S ribosomal subunit and restrain the extension of early polypeptide chains. Chloramphenicol binds to the 50S ribosomal subunit blocking peptidyl transferase reaction. Aminoglycosides inhibit initiation of protein synthesis and bind to the 30S ribosomal subunit (Leach *et al.,* 2007).

#### 1.8.3 Interference With Nucleic Acid Synthesis

Rifampicin interferes with a DNA-directed RNA polymerase. Quinolones inhibit DNA synthesis with interference of type II topoisomerase, DNA gyrase and type IV topoisomerase during replication cycle causing double strand break (Strohl, 1997).

#### 1.8.4 Inhibition Of A Metabolic Pathway

Sulfonamides (e.g. sulfamethoxazole) and trimethoprim each block the key steps in the folate synthesis, which is a cofactor in the biosynthesis of nucleotides, the building blocks of DNA and RNA (Strohl, 1997).

#### 1.8.5 Disorganizing Of The Cell Membrane

The primary site of action is the cytoplasmic membrane of Gram-positive bacteria, or the inner membrane of Gram-negative bacteria. It is hypothesized that polymyxins exert their inhibitory effects by increasing bacterial membrane permeability, causing leakage of bacterial content. The cyclic lipopeptide daptomycin shows fast bactericidal activity by binding to the cytoplasmic membrane in a calcium-dependent manner and oligomerizing in the membrane, leading to an efflux of potassium from the bacterial cell and cell death (Straus and Hancock, 2006).

### 1.9 Antibiotic Resistance Mechanism

Antibiotic resistance is the reduction in effectiveness and action of a drug, such as an antimicrobial in curing a disease or infection. Most commonly, the term dosage failure is used in the context of resistance that pathogens have ‘‘acquired’’, that is, resistance has evolved. When an organism is resistant to more than one drug, it is said to be multidrug-resistant (Fisher and Mobashery, 2010). Microbial strains may have different kinds of mechanisms used in resistance which has been shown figuratively in figure 1.

**Figure 1:**
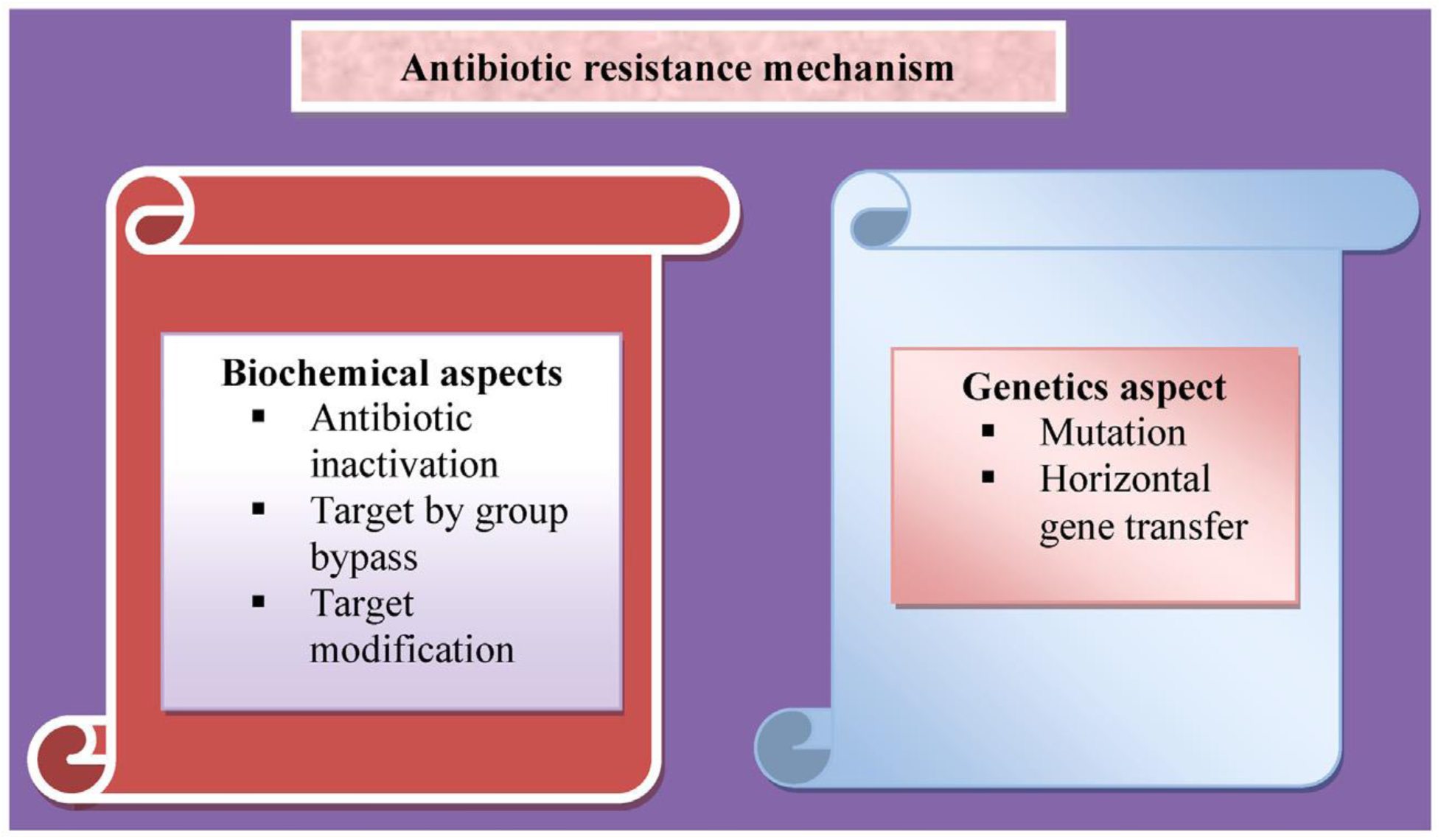
Biochemical and genetic aspects of antibiotic resistance mechanisms

#### 1.9.1 Antibiotic Inactivation

##### 1.9.1.1 By Hydrolysis

Many antibiotics have chemical bonds such as amides and esters which are hydrolytically susceptible. Several enzymes are known to destroy antibiotic activity by targeting and cleaving these bonds. These enzymes can often be excreted. Extended spectrum β-lactamases (ESBLs) mediate resistance to all penicillin, third generation cephalosporin (e.g. ceftazidime, cefotaxime, and ceftriaxone) and aztreonam, but not to cephamycin (cefoxitin and cefotetan) and carbapenems (Bonnet, 2004).

##### 1.9.1.2 By Redox Process

The pathogenic bacteria infrequently exploit oxidation or reduction of antibiotics. However, there are a few examples of these methods (Yang *et al.,* 2004). One of them is the oxidation of tetracycline antibiotics by the *TetX* enzyme (Bonnet, 2004).

#### 1.9.2 Antibiotic Inactivation By Group Transfer

The most diverse family of enzymes that show resistance is the group of transferases. These enzymes inactivate antibiotics (aminoglycosides, chloramphenicol, streptogramin, macrolides or rifampicin) by chemical replacement adenylyl, phosphoryl, or acetyl groups. The modified antibiotics are altered in their binding to a target. Chemical strategies include O-acetylation and N-acetylation (Blanchard, 2004; Schwarz *et al.,* 2004), O-phosphorylation (Matsuoka and Sasaki, 2004), O-nucleotidylation (Falagas *et al*., 2010), O-ribosylation, O-glycosylation, and thiol transfer. These covalent modification strategies all require a co-substrate such as ATP, acetyl-CoA, NAD+, UDP-glucose, or glutathione for their activity and consequently these processes are restricted to the cytoplasm.

#### 1.9.3 Antibiotic Resistance Via Target Modification

The second major mechanism for resistance is the modification of the antibiotic target site so that the antibiotic is not active enough to bind properly. However, it is possible for mutational changes to occur in the target that reduces the susceptibility to inhibition while retaining cellular function (Besier *et al.,* 2003).

#### 1.9.4 Antibiotic Resistance Via Mutations

There is a considerable number of biochemical mechanisms of bacterial antibiotic resistance. These are based on mutational occurrences, like the gene sequence, and coding for the target site of certain antibiotics (Ruiz, 2003). The variations in the expression of antibiotic uptake or of the efflux systems may also be modified by mutation (e.g. the reduced expression or absence of the OprD porin of *Pseudomonas aeruginosa* reduces the permeability of the cell wall to carbapenems) (Wolter *et al.,*2004).

#### 1.9.5 Antibiotic Resistance Via Horizontal Gene Transfer

A principal mechanism for the spread of antibiotic resistance is by exchange of hereditary material. Antibiotic resistance genes may be transferred by different mechanisms of conjugation, transformation, or transduction. Over the years, β-lactamase enzymes that have an extended spectrum of activity (ESBL) against most beta-lactams, including cephalosporins but not carbapenemases, have been discovered. One of these, CTX-M-15, initially found in *Escherichia coli* (Bush and Fisher, 2011; Woodford *et al.,* 2011). It is often found on highly mobile *IncFII* plasmids and associated with mobile genetic element *IS26*. The risk of infection and danger of disease is especially high in people with delayed hospitalization catheterization, nursing home residency, past anti-infection treatment, fundamental renal or liver pathology (Nordmann *et al.,* 2011).

### 1.10 Antibiotic Resistance Of Staphylococci

Antibiotics that are used against *Staphylococcus* spp. work in either of these ways, targeting cell wall synthesis, protein synthesis, nucleic acid synthesis, and other metabolic pathways. Selection of the antibiotics that are used in clinical and agricultural settings have promoted the evolution and spread of genes that confer resistance (Allen *et al.,* 2010). Resistance to various antibiotics can be either internal or acquired. Acquired by horizontal gene transfer, through various mobile genetic elements (such as plasmids, transposons, and integrons), enzymatic inactivation of the drug, and bypassing of the drug target. Internal mechanism includes mutational modification of gene targets, over expression of various efflux pumps. The exposure to antibiotics may lead to the formation of persister cells, small colony variants (SCVs), biofilms and increase efflux pump (Kwon *et al.,* 2008; Lewis, 2008).

### 1.11 Aim Of The Study

The aim of this study is to isolate DNase positive staphylococci from reading tables in halls of residence within the Obafemi Awolowo University, Ile-Ife, Osun State, Nigeria, and determine their antibiotics susceptibility pattern.

## CHAPTER TWO

## MATERIALS AND METHODS

### 2.1 MATERIALS

#### 2.1.1 EQUIPMENT

### The equipments used are: incubator, autoclave, Bunsen burner, refrigerator, light microscope, colorimeter, weighing balance, spatula, forceps, hot air oven, homogenizer, pipette filler, inoculating loops, inoculating needles

#### 2.1.2 GLASS WARES

The glass wares used include: conical flasks, pipettes, test tubes, glass slides, measuring cylinders.

#### 2.1.3 REAGENTS

They include: crystal violet, 70% ethanol, absolute (100%) ethanol, safranin, Gram’s iodine, hydrogen peroxide (H_2_O_2_) and immersion oil, distilled water and sterile water.

#### 2.1.4 MEDIA

They include: nutrient agar, nutrient broth, mannitol salt agar (MSA), Mueller-Hinton agar, DNase agar, normal saline. Other materials used were swab sticks, cryo-vails, aluminum foil, oxoid antibiotics disc, Petri dishes and cotton wool.

### 2.2 METHODS

#### 2.2.1 STERILIZATION OF MATERIALS

The following sterilization techniques were carried out during this research:

a. All inoculating loops and needles were flamed till red hot in Bunsen flame before use.
b. All glass wares used were sterilized in a hot air oven at 160^°^C for two hours before use.
c. The mouth tips of the conical flasks were constantly flamed to avoid contamination when pouring agar into different Petri dishes.
d. The glass slides were cleaned with 70% ethanol to ensure they were grease free and the workbench was disinfected with 70% ethanol before and after each experiment.

#### 2.2.2 PREPARATION AND STERILIZATION OF MEDIA

The media used were prepared according to the manufacturer’s instruction by weighing on a weighing balance the appropriate amount of the media in powdered form into an appropriate volume of distilled water in a conical flask and then homogenized on hot plate.

For solid media, the media in solution after homogenization was autoclaved at 121^°^C, 15psi for 15 minutes to allow for complete sterilization of the media and then allowed to cool to 45 ^°^C before pouring into Petri dishes. The media was allowed to cool and set before inverting the Petri dishes to prevent the condensate from dropping to the agar surface.

The liquid media was prepared by dissolving the media in powder in appropriate volume of water and homogenized, it was then dispensed into test tubes and autoclaved at 121^°^C, 15psi for 15minutes to allow for complete sterilization of the media and then allowed to cool.

#### 2.2.3 SAMPLE LOCATION

The samples were collected from tables of reading rooms at the following halls of residence in Obafemi Awolowo University, Ile-Ife, Osun State, Adekunle Fajuyi Hall, Ladoke Akintola Hall (Sports), Nnamdi Azikwe postgraduate Hall of residence, and Alumni Hall of Residence.

#### 2.2.4 SAMPLE COLLECTION

The samples were collected from the halls of residence using sterile swab sticks dipped into normal saline. Different parts of the reading tables were swabbed, and the swab sticks were taken immediately to the laboratory for processing.

#### 2.2.5 ISOLATION OF STAPHYLOCOCCI SPECIES

The swab sticks used in collection of samples were aseptically inoculated into prepared 3ml nutrient broth. The nutrient broth was then incubated at 37 ^°^C for 18-24 hours. The broth culture was then aseptically streaked on mannitol salt agar plates and incubated at 37 ^°^C for 48 hours. Distinct white or yellow, dome shaped colonies presumptively identified as staphylococci were again picked and inoculated into prepared 3ml of nutrient broth and incubated for 18-24 hours.

The culture broth was then streaked on nutrient agar, incubated at 37°C for 18-24 hours. The pure isolates were observed for their morphological characteristics on plates followed by identification and biochemical tests.

#### 2.2.6 IDENTIFICATION TESTS

##### 2.2.6.1 GRAM STAINING

###### Principle

Gram staining method is one of the most important procedures in Microbiology and was developed by Danish physician Hans Christian Gram in 1884. Gram staining is still the cornerstone of bacterial identification and taxonomic division, these method separate bacteria into groups of Gram positive and Gram negative, based on their cell wall composition. Gram positive bacteria appear purple under the microscope as they retain the crystal violet (primary stain) because of the thick peptidoglycan layer cell wall. The Gram-negative bacteria appear pink, because they have a thin peptidoglycan layer, so they lose the crystal violet stain and absorb the safranin stain (counter stain).

###### Procedure

Glass slide was cleaned with 70% ethanol to make it grease free, a drop of distilled water was placed on the slide using an inoculating loop. Two to three colonies of the 18-24 hours isolates were then picked from nutrient agar plates using a sterile inoculating loop to make a thin smear on the slide. The smear was allowed to air dry and was heat fixed by passing it over flame gently. The slide was then flooded with crystal violet (primary stain) and allowed to stay for 60seconds. It was then rinsed off gently with water running from a tap. The slide was flooded with Gram’s iodine for 45 seconds and then rinsed off with running water. After which the slide was flooded with safranin for 30 seconds and rinsed off under running water. The slide was then allowed to air dry and a drop of immersion oil was placed in the slide after which it was viewed under oil immersion (×100 magnification) lens using a light microscope.

##### 2.2.6.2 CATALASE TEST

###### Principle

Microorganisms that live and dwell in oxygenated environments can produce the enzyme catalase to neutralize toxic forms of oxygen metabolites, H_2_O_2._

Catalase breaks down Hydrogen peroxide (H_2_O_2_) into water and oxygen resulting in bubble formation.

###### Procedure

A drop of hydrogen peroxide was placed on a clean slide, a few isolate colonies were then transferred to the slide using a sterile wire loop. The rapid production of bubbles indicates a positive test while appearance of no bubbles indicates the organism inability to produce catalase.

##### 2.2.6.3 DNASE TEST

###### Principle

DNA Hydrolysis test or Deoxyribonuclease (DNase) test is used to determine the ability of an organism to hydrolyze DNA and utilize it as a source of carbon and energy for growth. An agar medium; DNase agar, a differential medium is used to perform this test.

###### Procedure

A line streak of a loopful from an 18-24 hours old culture on nutrient agar was made on the prepared DNase agar and incubated at 37 ^°^C for 18-24 hours. After incubation, the plates were flooded with one molar (1M) hydrochloric acid (HCl) and left for about 5minutes. The presence of a clear zone around the line of streak indicates a positive test for DNase reaction while the absence of a clear zone indicates a negative result.

##### 2.2.6.4 OXIDASE TEST

###### Principle

The oxidase test is used to identify and detect bacteria that produce cytochrome c oxidase, an enzyme of the bacteria electron transport chain. When present, the cytochrome c oxidase oxidizes the reagent (tetramethyl-p-phenylenediame) to indophenols which produce a purple colour on the oxidase strip as end product. When the enzyme is absent, the reagent remains reduced and no reaction on the strip and then it is colourless.

###### Procedure

On an oxidase paper strip, a few colonies of the pure isolates were picked with an inoculating loop and placed on the paper strip. After 2 minutes, it was observed for colour change. Presence of purple coloration indicates a positive test whereas the appearance of no coloration indicates a negative test.

##### 2.2.6.5 ANTIBIOTIC SUSCEPTIBILITY TEST

###### Principle

This test is used to determine the susceptibility of an organism to different antibiotics. Antibiotics are known to be produced partly or wholly from microorganisms or synthetically manufactured and kill or inhibit the microbial growth even in low concentration.

It is a quantitative assay that is done by placing antibiotics of a known concentration infused in paper disks on the surface of an already inoculated Mueller Hinton agar plate. The antibiotics diffuse from the disks into the agar creating a concentration gradient on the agar. An absence of growth around the disk (called zone of inhibition) is measured to determine if the organism is susceptible, intermediate and resistant to the antibiotic. Measured zones were interpreted according to the Clinical and Laboratory Standard Institute standards (CLSI) of 2016.

The antibiotics used were rifampicin (RD) 5µg, cefoxitin (FOX) 30µg, erythromycin (E) 15µg, clindamycin (DA) 2µg, chloramphenicol (C) 30µg, tetracycline (TE) 30µg and ciprofloxacin (CIP) 5µg.

###### Procedure

The colonies from the pure isolates were inoculated into normal saline and then standardized to 0.5 McFarland standard (1×10^8^cfu/ml). A sterile swab stick was then dipped into the tube containing the standardized organism and squeezed against the edge to drain excess fluid in the swab. The swab was then streaked on the Mueller-Hinton agar and the plate was allowed to dry for about 2 minutes. Each antibiotics disc used was then placed on the surface of the agar using sterile forceps. The inoculated plates were then inverted and incubated at 35 ^°^C for 18-24 hours. The zone of inhibition observed was measured for each antibiotic used and compared to the standard (CLSI,2016).

## 3.0 RESULTS

### 3.1 INFORMATION ON THE SAMPLES OBTAINED

A total of 23 isolates from 114 samples were obtained from reading rooms in selected halls of residence in Obafemi Awolowo University, Ile-Ife, Osun State, Nigeria.

### 3.2 IDENTIFICATION TEST

The staphylococci isolates were either white or yellow in colour, circular, small, mucoid, and raised with distinct colonies on mannitol salt agar plates. They were all Gram-positive cocci, catalase positive and oxidase negative. Twelve were DNase positive and eleven were DNase negative.

### 3.3 ANTIBIOTIC SUSCEPTIBILITY PATTERN

From the twelve DNase positive staphylococcal isolates, one was resistant to cefoxitin, rifampicin and clindamycin, two to erythromycin, four to chloramphenicol and five to tetracycline, all isolates were susceptible to clindamycin (Table 3).

**Table 3.1:** Table showing the antibiotic susceptibility pattern of the isolated DNase positive staphylococci on the tables of selected reading rooms in Obafemi Awolowo University, Ile-Ife, Osun State.

| ANTIBIOTICS | SUSCEPTIBLE [N/%] | INTERMEDIATE [N/%] | RESISTANT [N/%] |
| --- | --- | --- | --- |
| RIFAMPICIN 5 µg | 10 [84%] | 1 [8%] | 1 [8%] |
| CEFOXITIN 30 µg | 11 [92%] | 0 | 1 [8%] |
| ERYTHROMYCIN 15 µg | 6 [50%] | 4 [33%] | 2 [17%] |
| CLINDAMYCIN 2 µg | 9 [75%] | 2 [17%] | 1 [8%] |
| CHLORAMPHENICOL 30 µg | 8 [67%] | 0 | 4 [33%] |
| TETRACYCLINE 30 µg | 7 [58%] | 0 | 5 [42%] |
| CIPROFLOXACIN 5 µg | 12 [100%] | 0 | 0 |

### 3.4 ISOLATE LOCATION

The isolates location were the reading tables of Adekunle Fajuyi, Ladoke Akintola (Sports), Nnamdi Azikwe Postgraduate, and Alumni hall of residence within Obafemi Awolowo University.

**Table 3.2.**
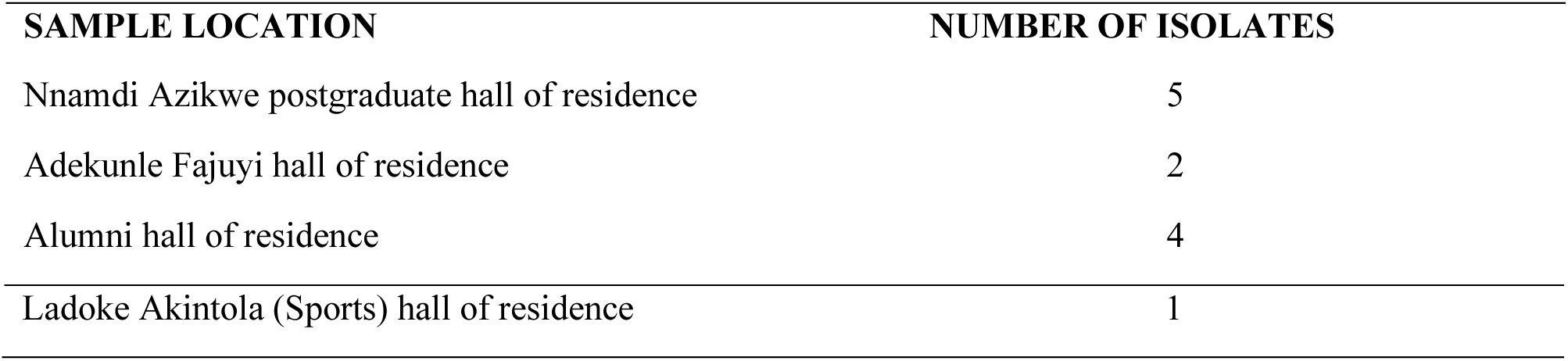
showing the isolates location and number of isolates from each location.

## 4. DISCUSSION AND CONCLUSION

Antimicrobial resistance is a global problem and threat to humanity because antibiotics-resistance bacterial infections are getting more difficult to treat and curb daily (Barbosa and Levy, 2000; Centre for disease control and prevention, 2013; Rodriguez *et al.,* 2013). Few studies have described the antimicrobial resistance profiles of staphylococci strains in the community, specifically in school environments. School vectors including humans and fomites (Uhlemann *et al.,* 2011; Mehta and Akul, 2011; Davis *et al.,* 2012; Fritz *et al.,* 2014). In this study, the total number of tables in reading halls swabbed (n = 68) and 116 samples collected, of which 26 were confirmed *Staphylococcus* and 12 were confirmed to be DNase positive.

One testing the susceptibility of the twelve DNase positive staphylococci to antibiotics, five (42%) were resistant to tetracycline, four (33%) to chloramphenicol, two (17%) to erythromycin and one each (8%) to rifampicin, cefoxitin, and clindamycin. Ciprofloxacin showed the best activity by having no isolate resistant to its action. This study reveals that reading tables serves as reservoir for microorganisms of which includes staphylococci species which accumulates from different sources, over long periods of time without proper cleaning. Tables can help transmit the staphylococci to other surfaces which may lead to infection, proper treatment of these infections with different classes of antibiotics have been encouraged, but antibiotics have different mode of action, the antibiotics of choice depends on the group of organisms causing the infection (Paul and Diana, 2006; Bairy *et al.,* 2013).

With this study, ciprofloxacin has shown the best activity on DNase positive staphylococci isolated from the reading tables, but should be used as the last resort, only if other antibiotics such as clindamycin, cefoxitin, erythromycin and rifampicin have not worked, in studies of DNase positive staphylococcal infections. Also, regular disinfection of tables and regular cleaning of reading rooms should be encouraged to reduce the risk of community acquired staphylococcal infections among students of the university.

## Data Availability

All data produced in the present work are contained in the manuscript

### APPENDIX I: MEDIA AND MEDIA COMPOSITION

**Mannitol salt agar (MSA)**

| Composition | gram/litre |
| --- | --- |
| Beef extract | 1.0 |
| Balanced peptone no.1 | 10.0 |
| Sodium Chloride | 75.0 |
| D-mannitol | 10.0 |
| Agar no.2 | 12.0 |
| Phenol Red | 0.025 |
| Final pH 7.4 ± 0.2 at 37°C |  |

To prepare this medium, 108g of dehydrated MSA powder was dissolved in 1000ml of distilled water and heated to homogenize the medium. This was the sterilized by autoclaving at 121°C (15psi) for 15minutes. It was then allowed to cool to 45°C and then poured into petri dishes and allowed to set. The plates were turned upside down to avoid condensation of moisture into the medium.

**Nutrient agar (NA)**

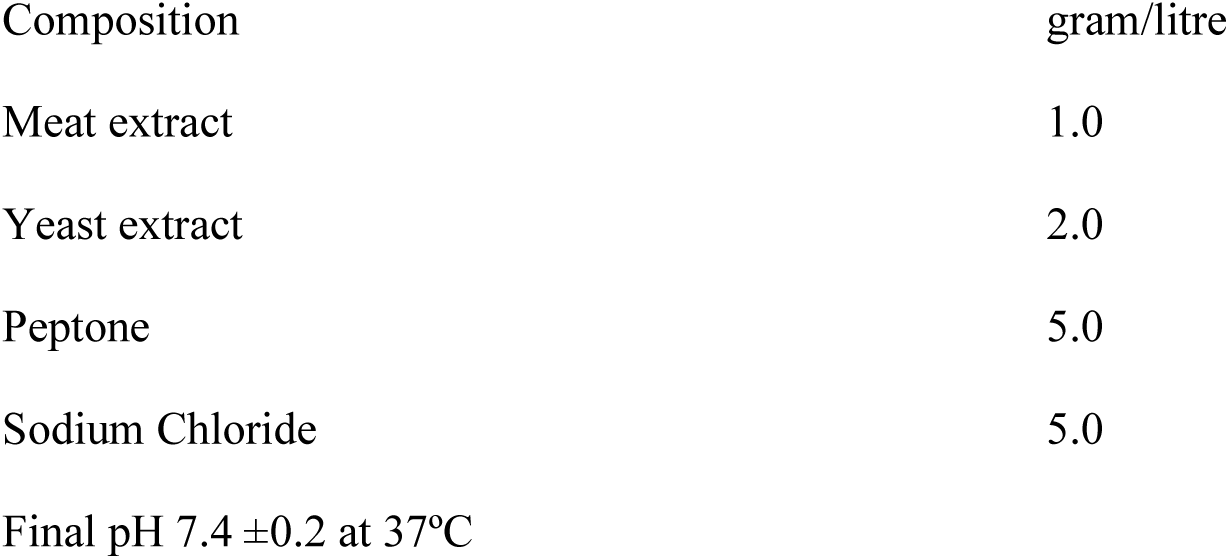

To prepare this medium, 28g of nutrient agar powder was weighed out and suspended in 1000ml of distilled water in a conical flask. This was the heated to homogenize it. It was then sterilized by autoclaving at 121°C for 15minutes. It was then allowed to cool to about 45°C, this was then dispensed into sterile petri dishes and allowed to set. The petri dishes were then inverted to avoid condensation of moisture into the medium.

**Mueller-Hinton agar (MHA)**

| Composition | gram/litre |
| --- | --- |
| Beef infusion solid | 4.0 |
| Starch | 1.5 |
| Casein hydroxylate | 17.5 |
| Agar | 15.0 |

Final pH 7.4 ±0.2 at 37°C

In preparing this, 38g of Mueller Hinton agar was suspended into 1000ml distilled water and boil to homogenize the medium completely. It was then cooled to about 45°C and then poured into plates.

**DNase agar**

| Composition | gram/litre |
| --- | --- |
| Enzymatic digest of casein | 15 |
| Enzymatic digest of animal tissue | 5 |
| Sodium Chloride | 5 |
| Deoxyribonucleic acid | 2 |
| Agar | 15 |

Final pH $7.3 \pm 0.2$ at 37°C

In preparing this media, 44g of DNase powder was suspended in 1000ml of distilled water, and was boiled to completely homogenize the medium. The desired amounts were dispensed into test tubes, plugged with cotton wool, and thereafter sterilized by autoclaving at 121°C for 15minutes. The medium was allowed to cool before use.

**Normal saline (0.85%)**

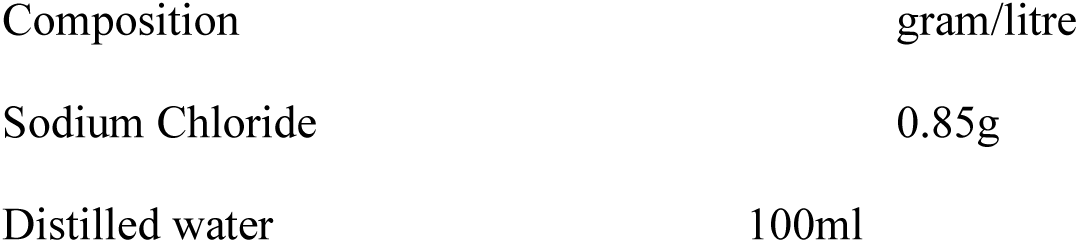

|  |  |
| --- | --- |
| Composition | gram/litre |
| Sodium Chloride | 0.85g |
| Distilled water | 100ml |

In preparing this, 0.85g of sodium chloride was dissolved in 1000ml of distilled water and was slightly heated to homogenise the medium, the desired amounts were dispensed into test tubes and plugged with cotton wool. The medium was sterilized by autoclaving at 121°C for 15minutes and allowed to cool before use.

### APPENDIX II:GRAM STAINING REAGENTS AND COMPOSITION

**Gram’s iodine solution**

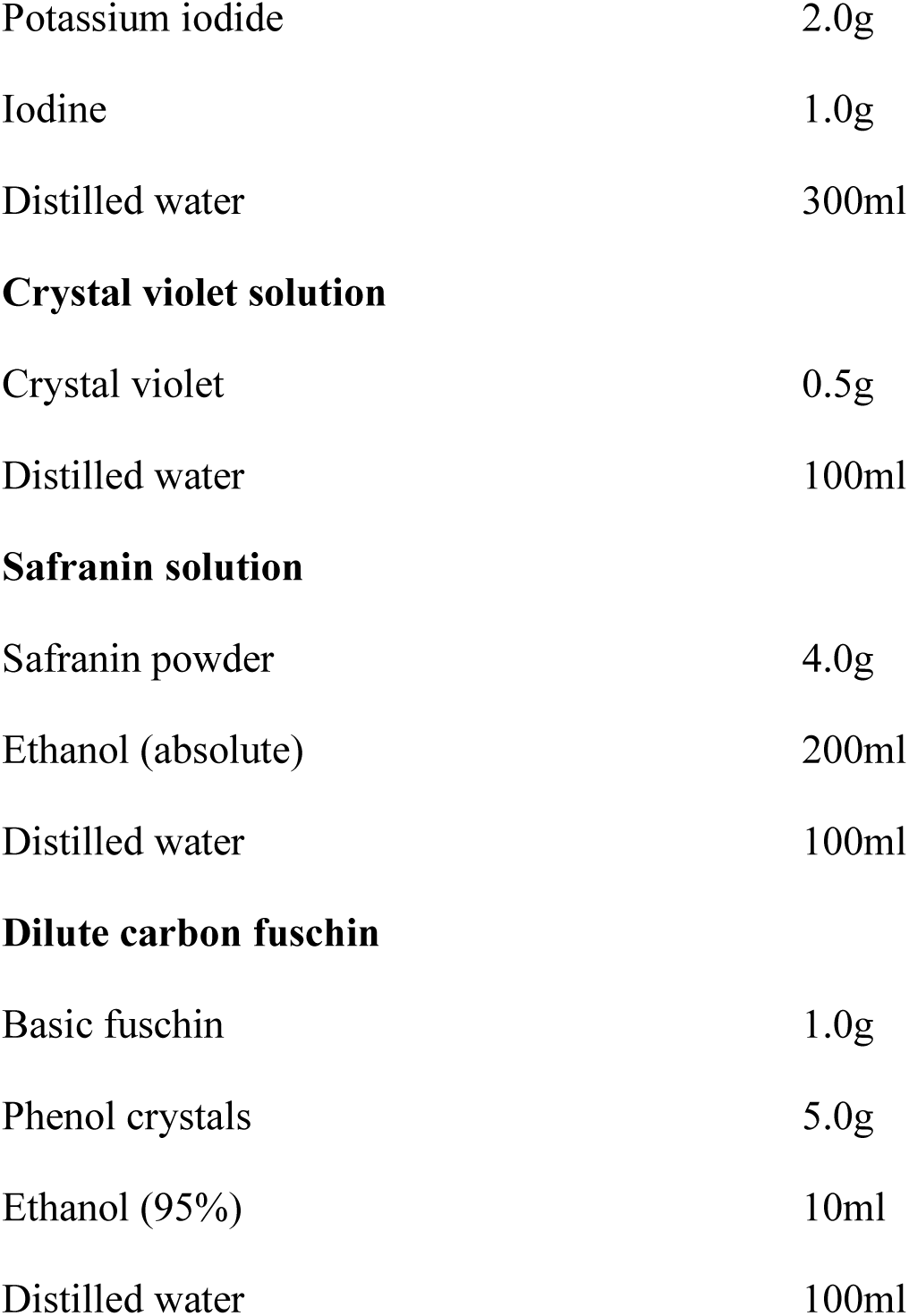

Gram's iodine solution
|  |  |
| --- | --- |
| Potassium iodide | 2.0g |
| Iodine | 1.0g |
| Distilled water | 300ml |
| Crystal violet solution |  |
| Crystal violet | 0.5g |
| Distilled water | 100ml |
| Safranin solution |  |
| Safranin powder | 4.0g |
| Ethanol (absolute) | 200ml |
| Distilled water | 100ml |
| Dilute carbon fuschin |  |
| Basic fuschin | 1.0g |
| Phenol crystals | 5.0g |
| Ethanol (95%) | 10ml |
| Distilled water | 100ml |

### APPENDIX III: MORPHOLOGICAL APPEARANCE OF THE ISOLATES

| S/N | ISOLATE CODE | APPEARANCE ON NUTRIENT AGAR | MANNITOL FERMENTATION | CATALASE | OXIDASE | GRAM STAINING |
| --- | --- | --- | --- | --- | --- | --- |
| 1 | PG 007B | White | Positive | Positive | Negative | +ve cocci |
| 2 | PG 009A | White | Positive | Positive | Negative | +ve cocci |
| 3 | PG 008A<sub>2</sub> | White | Positive | Positive | Negative | +ve cocci |
| 4 | PG 0064 | Cream | Positive | Positive | Negative | +ve cocci |
| 5 | PG 001B | Cream | Negative | Positive | Negative | +ve cocci |
| 6 | PG 004B | Cream | Negative | Positive | Negative | +ve cocci |
| 7 | PG 005A<sub>1</sub> | Cream | Positive | Positive | Negative | +ve cocci |
| 8 | PG 005B | Cream | Positive | Positive | Negative | +ve cocci |
| 9 | PG 001C | White | Positive | Positive | Negative | +ve cocci |
| 10 | PG 006C<sub>1</sub> | White | Positive | Positive | Negative | +ve cocci |
| 11 | ALM 004D<sub>1</sub> | Cream | Positive | Positive | Negative | +ve cocci |
| 12 | ALM 006B<sub>1</sub> | Yellow | Positive | Positive | Negative | +ve cocci |
| 13 | ALM 001C<sub>1</sub> | Yellow | Positive | Positive | Negative | +ve cocci |
| 14 | ALM 003D<sub>2</sub> | Yellow | Positive | Positive | Negative | +ve cocci |
| 15 | ALM 001<sub>1A</sub> | Cream | Positive | Positive | Negative | +ve cocci |
| 16 | ALM 005C<sub>3</sub> | Cream | Positive | Positive | Negative | +ve cocci |
| 17 | ALM 005B | Cream | Positive | Positive | Negative | +ve cocci |
| 18 | SPORT 001B<sub>1</sub> | White | Positive | Positive | Negative | +ve cocci |
| 19 | SPORT 003C | Cream | Negative | Positive | Negative | +ve cocci |
| 20 | FAJ 015A<sub>1A</sub> | White | Positive | Positive | Negative | +ve cocci |
| 21 | FAJ 002<sub>A2</sub> | White | Negative | Positive | Negative | +ve cocci |
| 22 | FAJ 001B<sub>3</sub> | Cream | Positive | Positive | Negative | +ve cocci |
| 23 | FAJ 006A | Cream | Positive | Positive | Negative | +ve cocci |
| 24 | FAJ 003A<sub>4</sub> | Cream | Negative | Positive | Negative | +ve cocci |
| 25 | FAJ 005B<sub>2</sub> | Cream | Negative | Positive | Negative | +ve cocci |
| 26 | FAJ 004B | Cream | Positive | Positive | Negative | +ve cocci |

### APPENDIX IV: TABLE OF ZONE OF INHIBITION OF DNASE POSITIVE STAPHYLOCOCCI

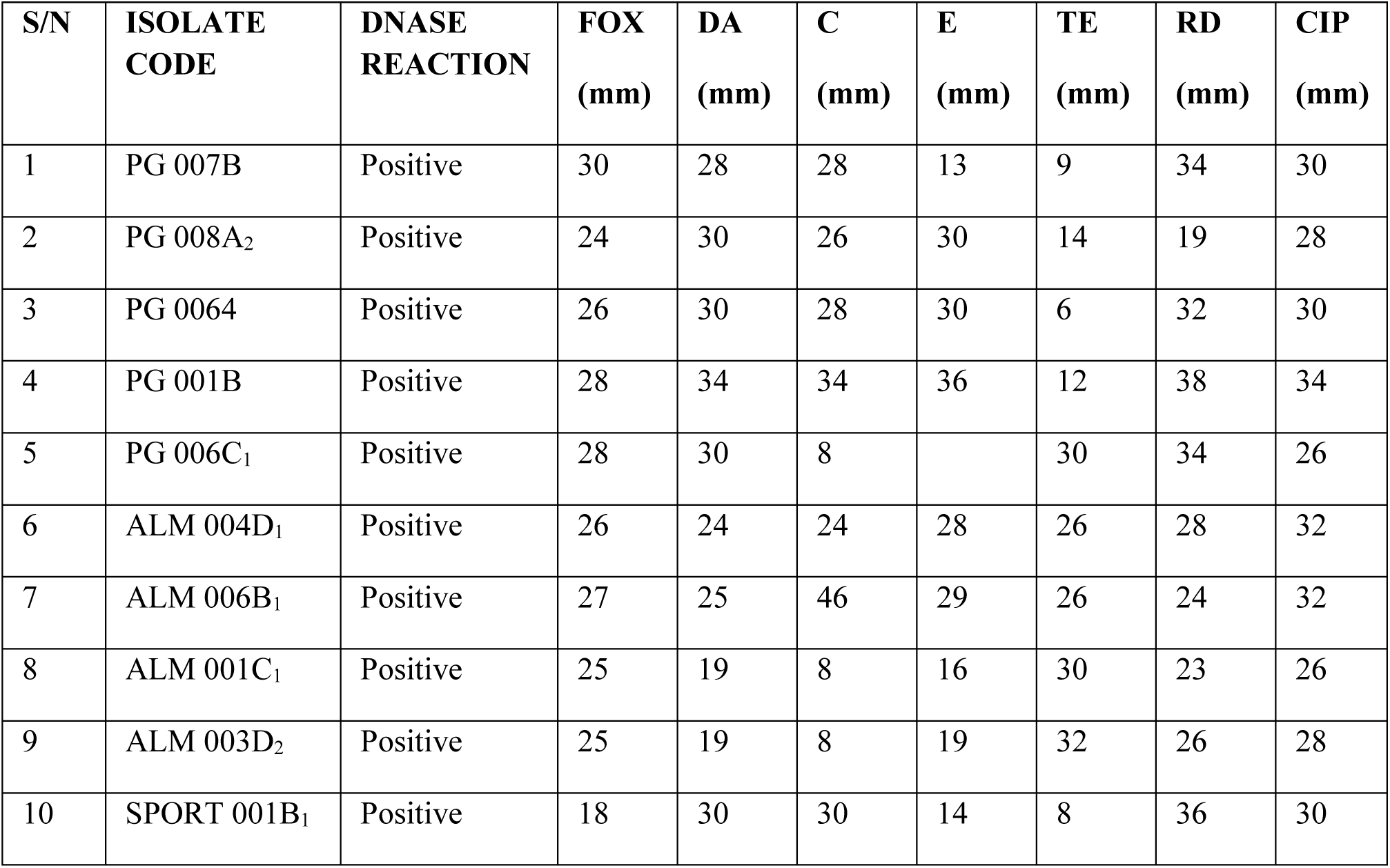
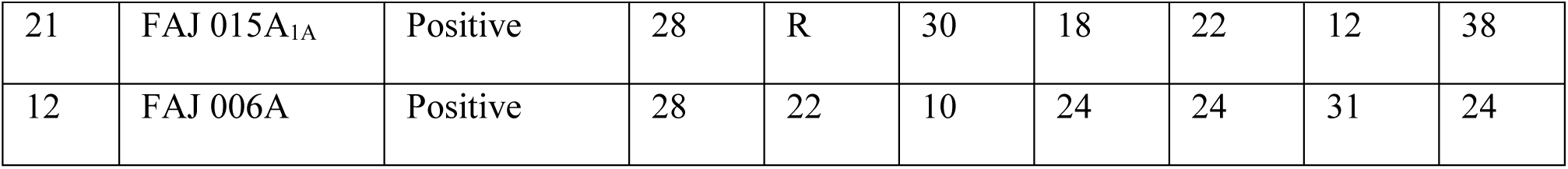

### APPENDIX V: TABLE OF CSLI STANDARD

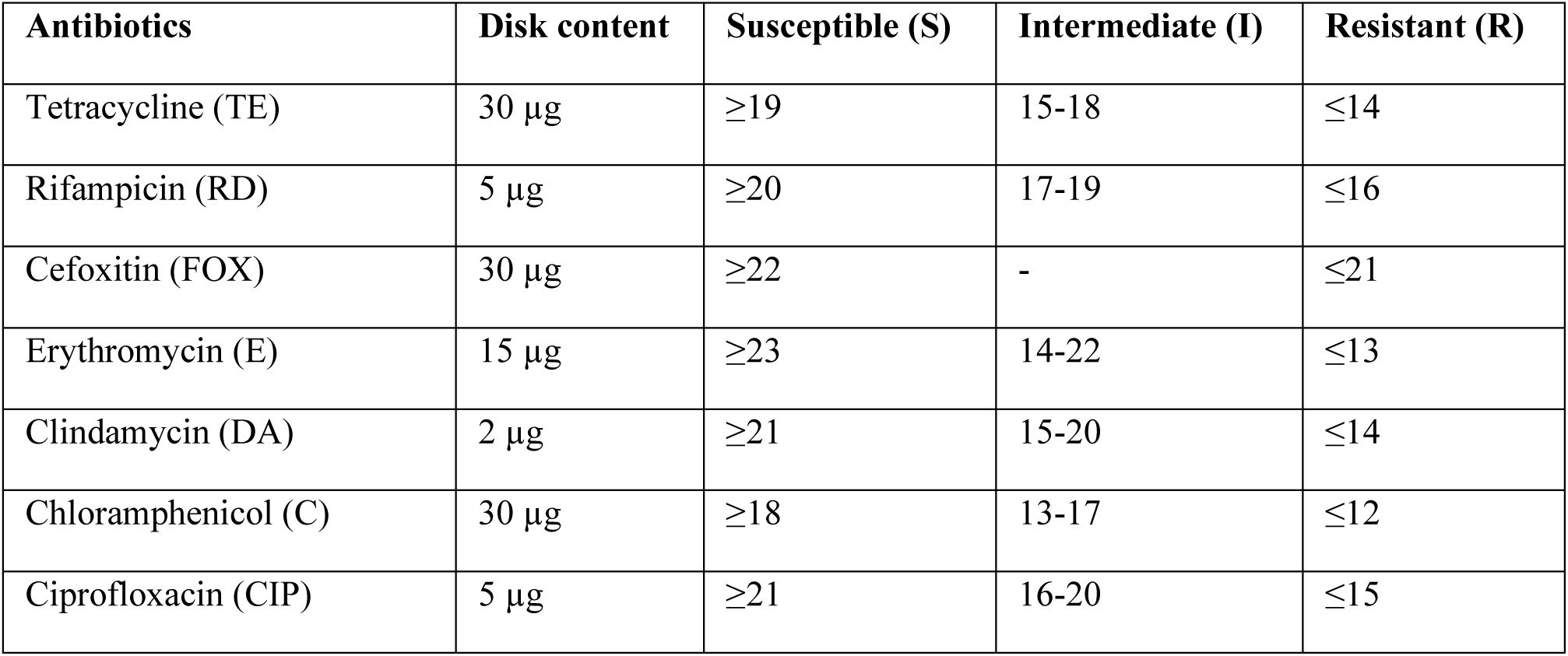

| Antibiotics | Disk content | Susceptible (S) | Intermediate (I) | Resistant (R) |
| --- | --- | --- | --- | --- |
| Tetracycline (TE) | 30 µg | ≥19 | 15-18 | ≤14 |
| Rifampicin (RD) | 5 µg | ≥20 | 17-19 | ≤16 |
| Cefoxitin (FOX) | 30 µg | ≥22 | - | ≤21 |
| Erythromycin (E) | 15 µg | ≥23 | 14-22 | ≤13 |
| Clindamycin (DA) | 2 µg | ≥21 | 15-20 | ≤14 |
| Chloramphenicol (C) | 30 µg | ≥18 | 13-17 | ≤12 |
| Ciprofloxacin (CIP) | 5 µg | ≥21 | 16-20 | ≤15 |

### APPENDIX V: TABLE OF SUSCEPTIBILITY PATTERN OF DNASE POSITIVE STAPHYLOCOCCI

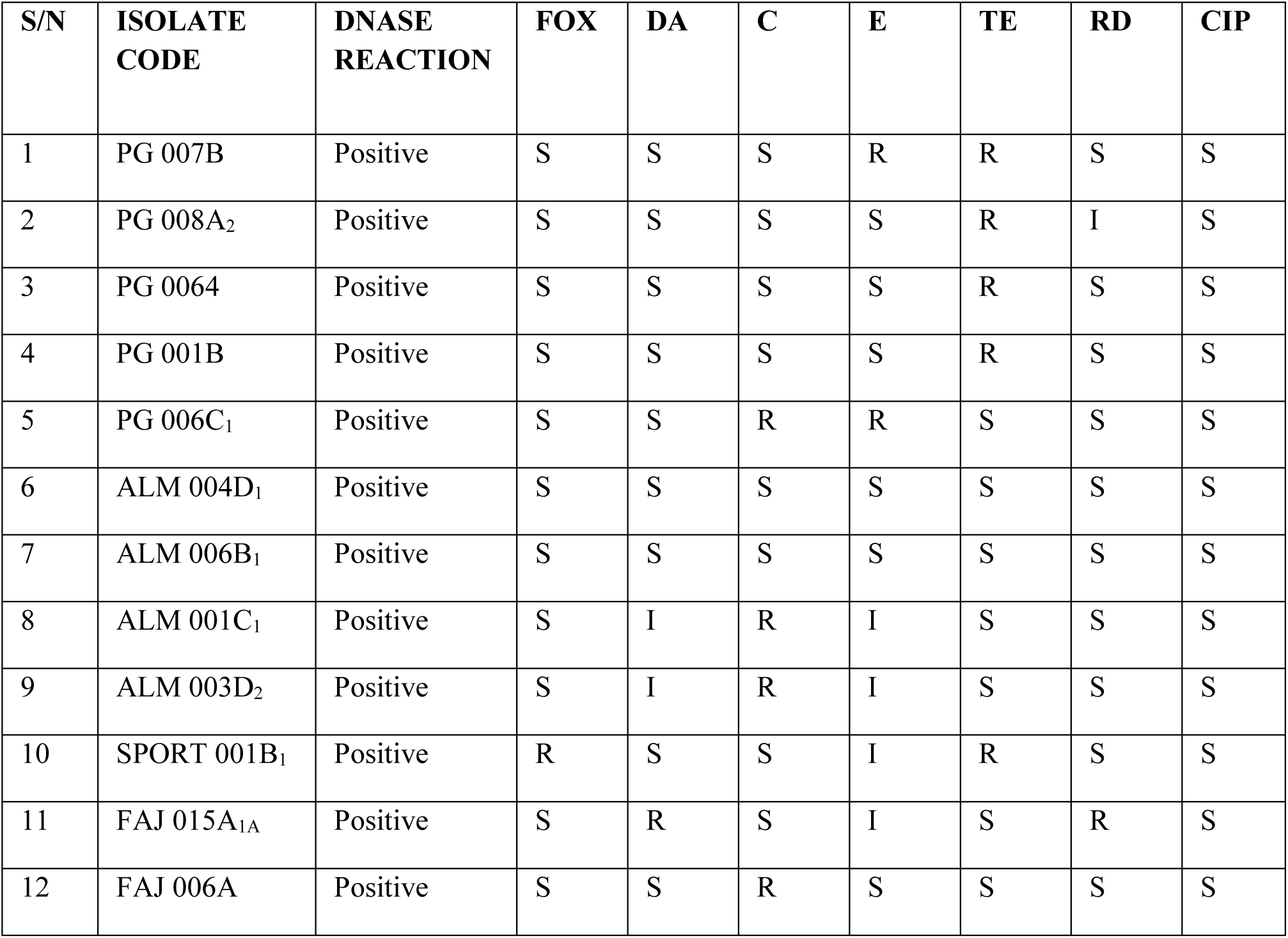

